# Impact of patient habitus and acquisition protocol on iodine quantification in dual source photon-counting CT

**DOI:** 10.1101/2022.12.16.22283594

**Authors:** Rizza Pua, Leening P. Liu, Michael Dieckmeyer, Nadav Shapira, Pooyan Sahbaee, Grace J. Gang, Harold I. Litt, Peter B. Noël

## Abstract

**Objective:** Evaluation of iodine quantification accuracy with varying iterative reconstruction level, patient habitus, and acquisition mode on a first-generation dual-source photon-counting computed tomography (PCCT) system.

**Methods:** A multi-energy CT phantom (20 cm diameter/small) was imaged with and without an extension ring (30 by 40 cm/large). It was equipped with various iodine inserts (0.2, 0.5, 1.0, 2.0, 5.0, 10.0, 15.0 mg/ml) and scanned over a range of radiation dose levels (CTDIvol 0.5, 0.8, 1.2, 1.6, 2.0, 4.0, 6.0, 10.0, 15.0 mGy) using four different acquisition modes: single source 120 kVp (SS120), 140 kVp (SS140) and dual-source 120 kVp (DS120), 140 kVp (DS140). Iodine density maps were produced with different levels of iterative reconstruction (QIR 0, 2, 4). To assess the agreement between nominal and measured iodine concentrations, root mean square error (RMSE) and Bland-Altman plots were generated by grouping different radiation dose levels (ultra-low: < 1.5 mGy; low: 1.5 – 5 mGy; medium: 5 – 15 mGy) and iodine concentrations (low: < 5 mg/ml; high: 5 – 15 mg/ml).

**Results:** Overall, quantification of iodine concentrations was accurate and reliable even at ultra-low radiation dose levels. With low and high iodine concentrations, RMSE ranged from 0.25 to 0.37, 0.20 to 0.38, and 0.25 to 0.37 mg/ml for ultra-low, low, and medium radiation dose levels, respectively. Similarly, for the three acquisition modes (SS120, SS140, DS 120, DS140), RMSE was stable at 0.31, 0.28, 0.33 and 0.30 mg/ml, respectively. Considering all levels of radiation dose, acquisition mode, and iodine concentration, the accuracy of iodine quantification was higher for the phantom without extension ring (RMSE 0.21 mg/ml) and did not vary across different levels of iterative reconstruction.

**Conclusions:** The first-generation PCCT allows for accurate iodine quantification over a wide range of iodine concentrations and radiation dose levels. Even very small concentrations of iodine can be quantified accurately at different simulated patient sizes. Stable accuracy across iterative reconstruction levels may allow further radiation exposure reductions without affecting quantitative results.

**Summary:** Clinical photon-counting CT provides excellent iodine quantification performance for a wide range of parameters (patient habitus, acquisition parameters, and iterative reconstruction modes) due to its excellent ultra-low dose performance.

**Key Results:** First-generation PCCTs are capable of accurately quantifying iodine over a wide range of radiation dose levels and iodine concentrations.

Further radiation exposure reductions may be possible given stable accuracy across iterative reconstruction levels.

In the future, accurate and precise iodine quantification will allow for the development of spectral-based biomarkers.

## Introduction

Intravenous administration of iodinated contrast during CT acquisitions can improve diagnostic utility for many clinical indications by improving contrast between structures with similar attenuation on non-contrast (native) scans. In addition to visual assessment of contrast enhancement, iodine measurements provide the ability to assess contrast uptake of various tissues, organs, and lesions quantitatively. However, CT systems are only capable of assessing the amount of iodine at best in a semi-quantitative way as highly attenuating materials, such as calcifications or bone, can have HU values in the same range as iodine. With beam hardening artifacts present in conventional CT, these materials can make it difficult for contrast-enhancing features to be differentiated from non-enhancing features.

Spectral CT can overcome challenges of conventional CT, such as beam hardening or non-quantitative measurements, in order to accurately determine iodine levels using iodine density maps. The latest addition to the family of spectral CT instruments is photon-counting computed tomography (PCCT)^1^. Many years of preclinical research have demonstrated superiority of photon-counting detectors (PCDs) over energy-integrating detectors (EIDs) in image quality (high contrast-to-noise ratio, high spatial resolution, low electronic noise, and reduced artifacts), quantitative imaging, and dose efficiency^2–8^. Since PCCT has made the transition into the clinical routine^9^, iodine quantification studies on first-generation clinical systems have further confirmed its superiority over EID-CT in applications like abdominal^10–12^, and cardiac imaging^13–15^.

Accurate iodine quantification is needed to ensure reliable clinical results. Consequently, the purpose of this study was to characterize iodine quantification performance of a first-generation clinical dual-source PCCT system (NAEOTOM Alpha, Siemens Healthineers). A phantom containing iodine inserts of various concentrations was utilized to investigate and evaluate the effects of patient habitus, acquisition mode, radiation dose, and iterative reconstruction on iodine quantification.

## Methods

### CT Phantom

A multi-energy CT phantom (Multi-energy CT, Sun Nuclear) (Figure 1) was used with and without an extension ring to simulate two patient sizes (20-cm diameter (‘small’) and 30 × 40 cm (‘large’)). The phantom contained varying concentrations of iodine (0.2, 0.5, 1, 2, 5, 10, and15 mg/ml), blood with iodine (blood + 2 mg/ml iodine and blood + 4 mg/ml iodine), blood, and brain tissue-equivalent inserts. In this study, analysis of iodine measurements was limited to 0.2 - 15 mg/ml iodine inserts. Ground truth iodine densities were based on information provided by the manufacturer. The arrangement of iodine inserts in the phantom is displayed in Figure 1.

**Figure 1.**
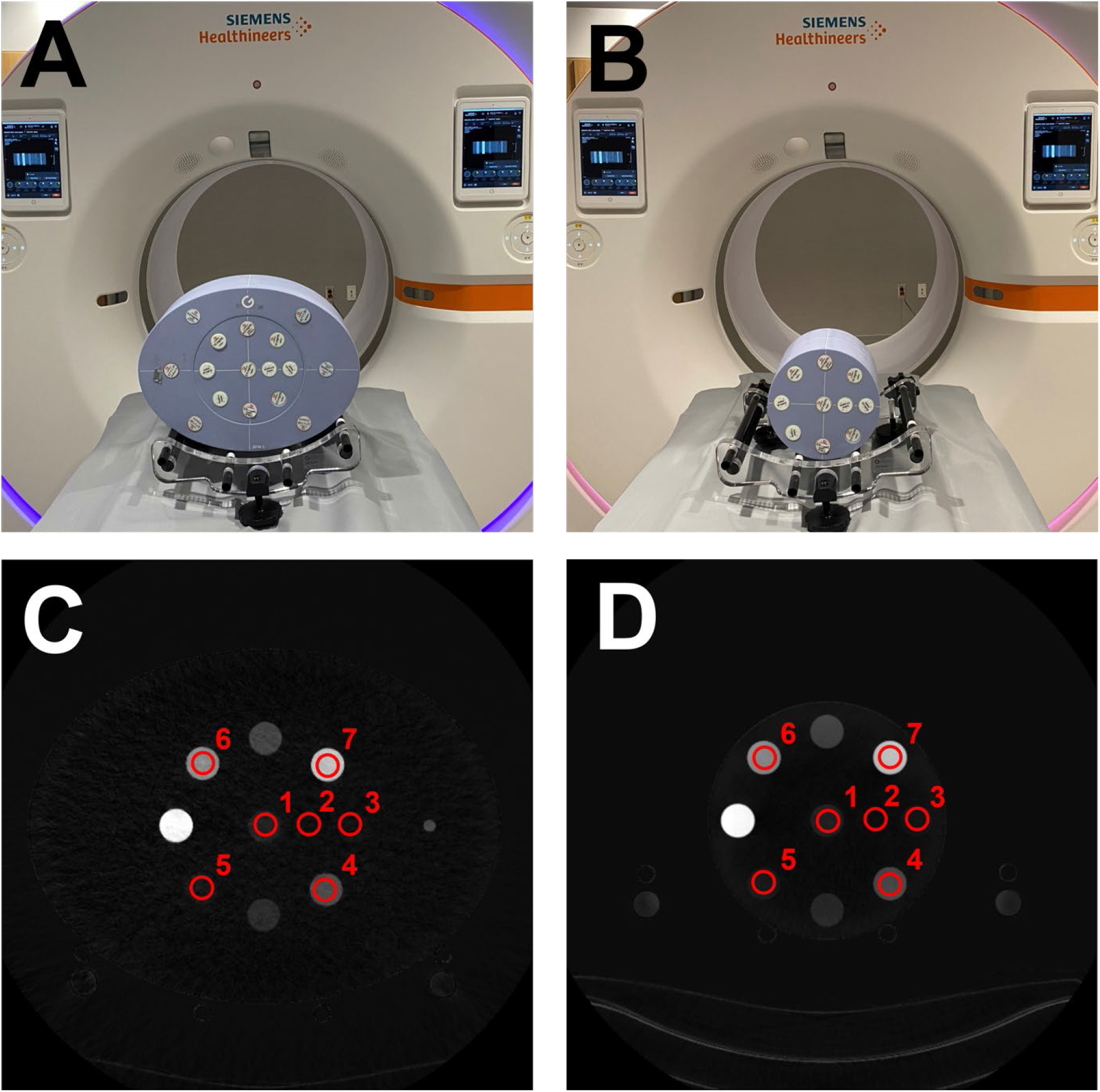
Experimental setup. (**A, B**) Photography of NAEOTOM Alpha with multi-energy CT phantom in large and small configurations. (**C, D**) Reconstructed iodine map slice of large and small phantom configurations with numbered iodine inserts of varying concentrations: 1, iodine 2 mg/ml; 2, iodine 0.2 mg/ml; 3, iodine 0.5 mg/ml; 4, iodine 5 mg/ml; 5, iodine 1 mg/ml; 6, iodine 10 mg/ml; 7, iodine 15 mg/ml. Iodine density maps are obtained from a single source 120 kVp acquisition mode at 10 mGy and reconstructed using QIR level 4. (WW,WL) = (22, 10) mg/ml.

### Image Acquisition & Reconstruction

All image acquisitions were performed on a first-generation clinical DS-PCCT in two acquisition modes: single-source (SS) mode and DS mode each at tube voltages of 120 and 140 kVp. For both configurations (small / large), the phantom was placed in the iso-center of the scanner (Figure 1) and a standard abdominal protocol was implemented for data acquisition and image reconstruction (Table 1). Without using any exposure modulations, scans were performed from ultra-low to medium radiation dose levels: CT dose index (CTDI_vol_) of 0.6, 0.8, 1.2, 1.6, 2, 4, 6, 10, and 15 mGy. The CTDI_vol_ range is equivalent to an effective dose range between 0.54 mSv to 13.50 mSv (k = 0.015 mSv mGy^-1^ cm^-1^) for an abdomen scan with a length of 60 cm. Based on the size of the patient, the selected dose range represents the clinical range from ultra-low doses to standard doses. Each was repeated three times without moving the phantom in-between scans. Individual scans were reconstructed with three levels of quantum iterative reconstruction (QIR) (QIR0 = iterative reconstruction turned off, QIR2, QIR4) in spectral mode to generate iodine density maps. Additional 70 keV virtual monoenergetic images (VMIs) were reconstructed of the 6.0 mGy scan for each phantom size to automate region of interest (ROI) placement.

**Table 1.** Acquisition and reconstruction parameters.

|  |  |
| --- | --- |
| Scanner model | NAEOTOM Alpha |
| Acquisition mode | single-source, dual-source |
| Tube voltage | 120, 140 kVp |
| Rotation time | 0.25 |
| Spiral pitch factor | 1 |
| Collimation | 144 × 0.4 mm |
| Slice thickness | 3 mm |
| Iterative reconstruction | QIR0, QIR2, QIR4 |
| Reconstruction filter | Qr48 |
| Reconstructed field of view | 450 mm |
| Matrix size | 512 x 512 |
| Pixel spacing (in x and y) | 0.88 mm |

### Image Analysis

ROIs were prescribed on selected iodine-containing inserts on a 6.0 mGy VMI 70 keV for each phantom size and combination of acquisition parameters. These ROIs were copied to the iodine density images at other dose levels from the corresponding phantom size and combination of acquisition parameters. Mean and standard deviation were calculated across a total of 30 slices (3 repeated scans x 10 consecutive central slices) for each insert at each dose level. Measured iodine density corresponding to 0.2, 0.5, 1, 2, 5, 10, and 15 mg/ml iodine concentrations were plotted against expected values, and error bars depict the standard deviation (*σ*) of the mean. Within each iodine insert column, measured iodine density from different phantom sizes (small / large) and tube combinations (SS120, SS140, DS120, DS140) were arranged by increasing radiation dose groups (ultra-low: 0.6, 0.8, 1.2 mGy, low: 1.6, 2, 4 mGy, and medium: 6, 10, 15 mGy). The corresponding root-mean-square error (RMSE) was calculated and plotted separately.

The mean bias and measured bias between measured iodine density concentrations and expected values for each phantom size (small / large) and tube combinations (SS120, SS140, DS120, and DS140) over low radiation dose levels (CTDIvol = 1.6, 2, 4 mGy) were illustrated using a Bland-Altman (BA) plot. The mean bias and measured bias were separately calculated and plotted for low iodine concentrations (0.2, 0.5, 1, 2 mg/ml) and high iodine concentrations (5, 10, 15 mg/ml). Limits of agreement (LOA) were calculated as mean bias ±1.96 x (σ of bias). Corresponding results for ultra-low (CTDI_vol_ = 0.6, 0.8, 1.2 mGy) and medium (CTDI_vol_ = 6, 10, 15 mGy) radiation dose levels were summarized in separate tables.

To demonstrate the effect of QIR on iodine quantification, RMSE values were calculated over all iodine concentrations (0.2 - 15 mg/ml) for each QIR level (QIR0, QIR2, and QIR4) across all radiation dose levels (CTDI_vol_ = 0.6 - 15 mGy) acquired using SS120 acquisition mode. Additional RMSE values were separately calculated over low iodine concentrations (0.2, 0.5, 1, 2 mg/ml) and high iodine concentrations (5, 10, 15 mg/ml). The RMSEs were shown in three different plots labeled as all, low, and high. In each CTDI_vol_ column, RMSE values were arranged by increasing QIR levels (QIR0, QIR2, and QIR4).

Similar RMSE calculations were made to demonstrate the effect of patient habitus on iodine quantification. RMSE values were calculated over all iodine concentrations (0.2 - 15 mg/ml) for each phantom size (small / large) across all radiation dose levels (CTDI_vol_ = 0.6 - 15 mGy) acquired using SS120 acquisition mode. Moreover, RMSE values were separately calculated over low iodine concentrations (0.2, 0.5, 1, 2 mg/ml) and high iodine concentrations (5, 10, 15 mg/ml) resulting in three different plots labeled as all, low, and high.

## Results

Measured iodine concentrations and corresponding RMSE values are summarized in Figure 2. Overall, there was consistent iodine quantification accuracy across all iodine inserts between measured and reference values. For iodine concentrations 0.2 - 2 mg/ml, the RMSE values were below 0.3 mg/ml. RMSE values of less than 0.7 mg/ml were observed for 0.5 and 2 mg/ml iodine inserts for large phantom obtained using DS120 and DS140 tube combinations at ultra-low radiation dose levels. The majority of the RMSE values for 5 mg/ml were below 0.4 mg/ml. For 10 mg/ml iodine, the large phantom showed higher RMSE values ranging from 0.3 - 0.7 mg/ml compared to 0.1 - 0.3 mg/ml for the small phantom. The 15 mg/ml iodine insert demonstrated RMSE values less than 0.8 mg/ml except for an outlier of less than 1.15 mg/ml noted for the large phantom obtained using DS120 at ultra-low radiation dose levels.

**Figure 2.**
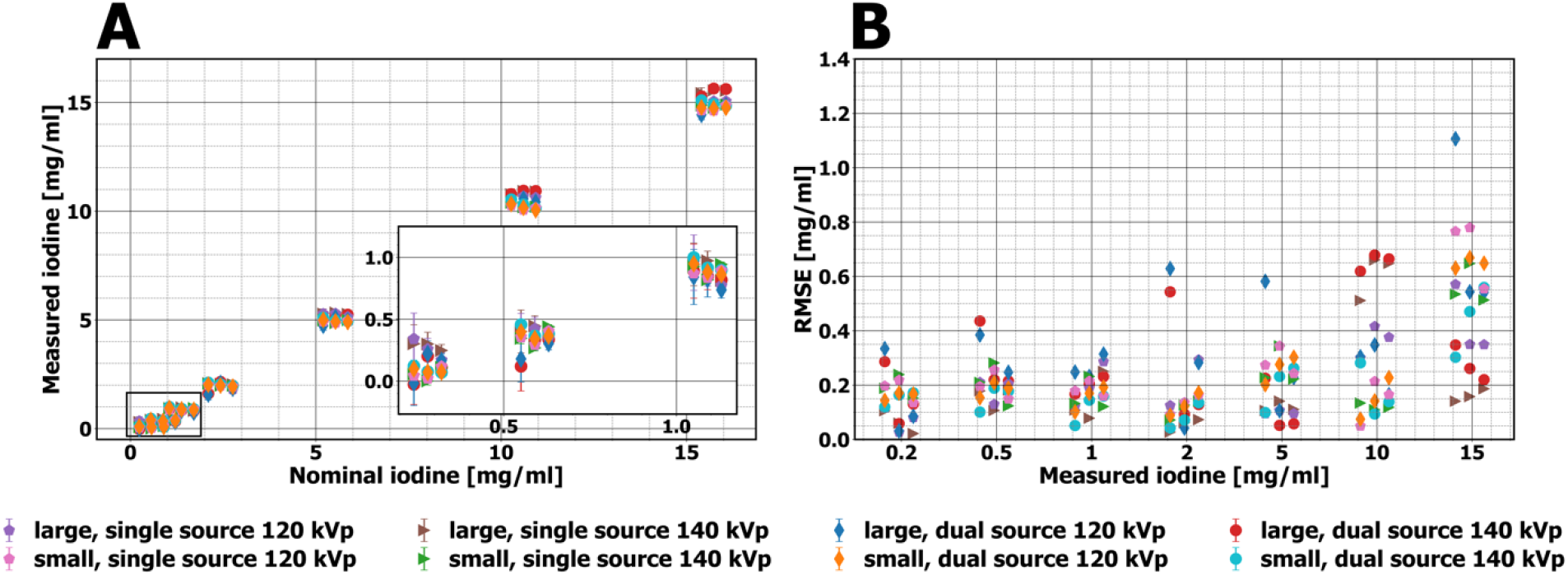
Comparison of (**A**) measured iodine densities [mg/ml] and (**B**) corresponding RMSEs [mg/ml] for individual inserts, ordered by increasing iodine concentration (0.2 mg/ml – 15 mg/ml). Within each column, measurements are ordered by increasing CTDI_vol_ groups (ultra-low: 0.6, 0.8, 1.2 mGy, low: 1.6, 2, 4 mGy, and medium: 6, 10, 15 mGy)). Phantom size (small / large) and acquisition parameters (SS120, SS140, DS120, and DS140) are coded in different colors and marker symbols.

Figures 3 and 4 illustrate the Bland-Altman plots for low and high iodine concentrations, respectively, separated for each phantom size (small / large) and tube combinations (SS120, SS140, DS120, and DS140) over low radiation dose levels (CTDI_vol_ = 1.6, 2, 4 mGy). In Figure 3, the mean bias measured for large and small phantom configurations at various tube combinations ranged from -0.11 mg/ml - -0.04 mg/ml and -0.22 mg/ml - -0.14 mg/ml, respectively. In Figure 4, the corresponding mean bias ranges were -0.11 mg/ml - 0.32 mg/ml and -0.44 mg/ml - -0.22 mg/ml for large and small phantoms, respectively. However, only a small difference between individual mean biases was observed across acquisition parameters for both phantom configurations. All bias measurements were within the calculated limits of agreement (LOA). For low iodine concentrations (Figure 3), the widths of the LOA from different acquisition modes ranged from 0.24 mg/ml - 0.44 mg/ml and 0.13 mg/ml - 0.23 mg/ml for the large and small phantoms, respectively. For high iodine concentrations (Figure 4), the corresponding LOA widths ranged from 0.94 mg/ml - 1.34 mg/ml and 0.8 mg/ml - 0.95 mg/ml. Bland-Altman results over ultra-low and medium radiation dose levels are summarized in Tables 2-3 and Tables 4-5, respectively.

**Table 2.** Bland-Altman results calculated for low iodine concentrations (0.2, 0.5, 1, 2 mg/ml), phantom sizes (small / large), and tube combinations (SS120, SS140, DS120, and DS140) over ultra-low radiation dose levels (CTDI_vol_ = 0.6, 0.8, 1.2 mGy).

| Phantom size | Acquisition mode | Bias [mg/ml] |  |  |  | Mean bias [mg/ml] | Limits of agreement |  |
| --- | --- | --- | --- | --- | --- | --- | --- | --- |
|  |  | Iodine 0.2 mg/ml | Iodine 0.5 mg/ml | Iodine 1 mg/ml | Iodine 2 mg/ml |  | Lower [mg/ml] | Upper [mg/ml] |
| Large | SS120 | 0.11 | -0.19 | -0.1 | -0.08 | -0.06 | -0.27 | 0.14 |
|  | SS140 | 0.06 | -0.15 | -0.11 | -0.01 | -0.05 | -0.21 | 0.11 |
|  | DS120 | -0.26 | -0.37 | -0.21 | -0.53 | -0.34 | -0.58 | -0.1 |
|  | DS140 | -0.27 | -0.43 | -0.16 | -0.49 | -0.34 | -0.59 | -0.08 |
| Small | SS120 | -0.19 | -0.19 | -0.18 | -0.09 | -0.16 | -0.25 | -0.08 |
|  | SS140 | -0.18 | -0.21 | -0.13 | -0.04 | -0.14 | -0.27 | -0.01 |
|  | DS120 | -0.14 | -0.15 | -0.1 | -0.08 | -0.12 | -0.18 | -0.06 |
|  | DS140 | -0.12 | -0.09 | -0.05 | 0.02 | -0.06 | -0.16 | 0.04 |

**Figure 3.**
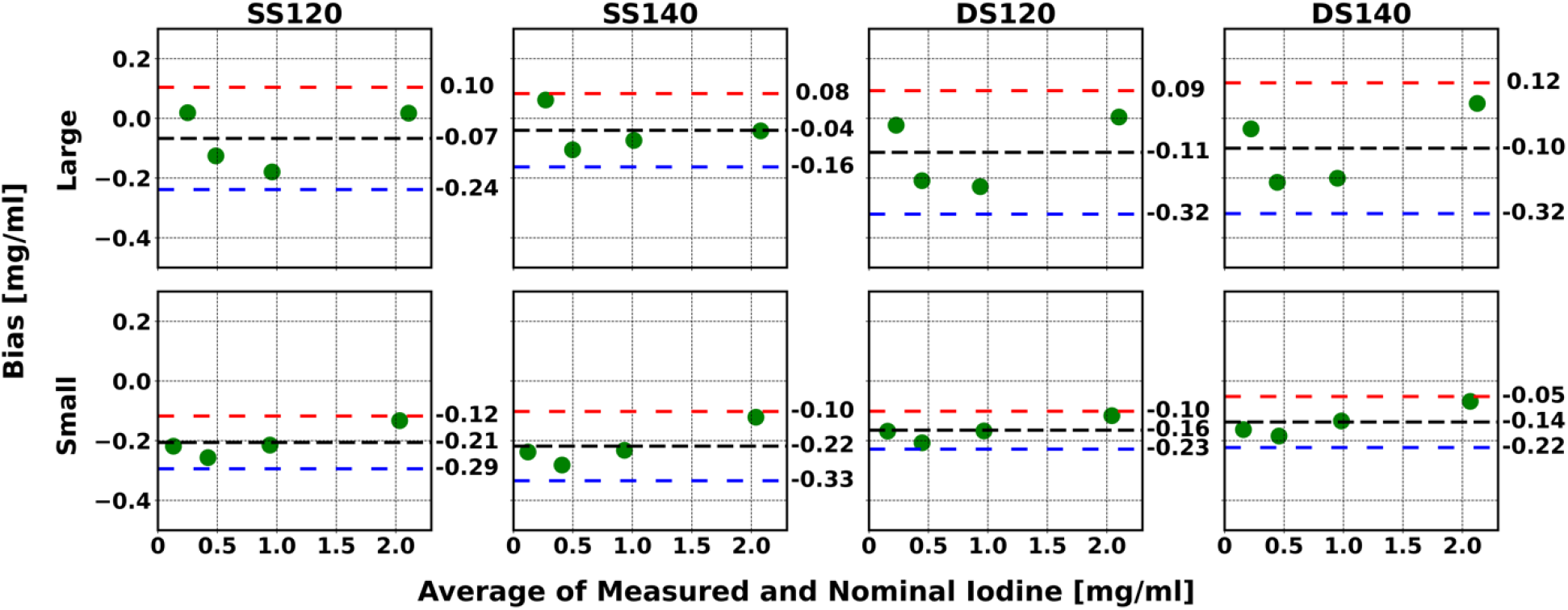
Bland-Altman plots illustrating the measured bias [mg/ml] and mean bias [mg/ml] between measured and nominal iodine concentrations versus the average of measured and nominal iodine concentrations [mg/ml] acquired for low iodine concentrations (0.2, 0.5, 1, 2 mg/ml), phantom sizes (small / large), and tube combinations (SS120, SS140, DS120, and DS140) over low radiation dose levels (CTDI_vol_ = 1.6, 2, 4 mGy). The black dashed line represents the mean bias. The red and blue dashed lines represent the upper and lower limits of the mean bias (from ±1.96σ).

**Figure 4.**
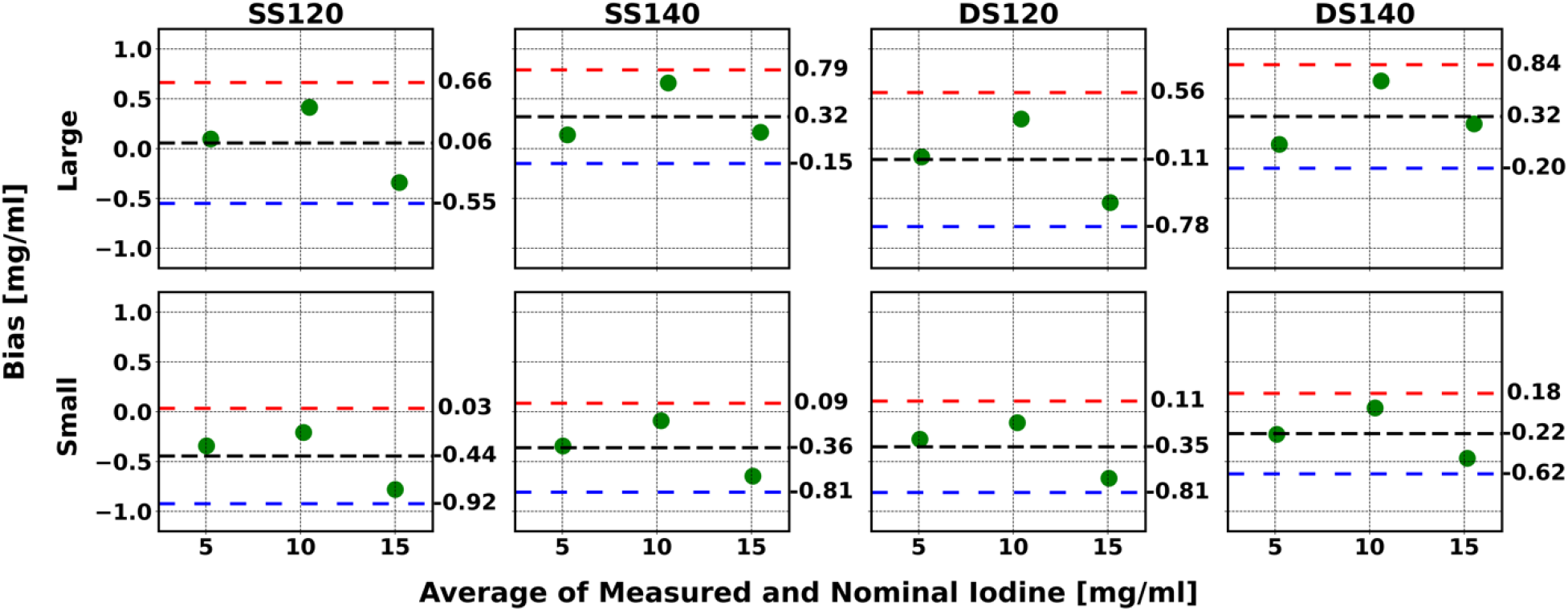
Bland-Altman plots illustrating the measured bias [mg/ml] and mean bias [mg/ml] between measured and nominal iodine concentrations versus the average of measured and nominal iodine concentrations [mg/ml] acquired for high iodine concentrations (5, 10, 15 mg/ml), phantom sizes (small / large), and tube combinations (SS120, SS140, DS120, and DS140) over low radiation dose levels (CTDI_vol_ = 1.6, 2, 4 mGy). The black dashed line represents the mean bias The red and blue dashed lines represent the upper and lower limits of the mean bias (from ±1.96σ).

The highest mean bias (−0.44 mg/ml) for ultra-low radiation dose levels occurred for the combination of the large phantom, DS120, and high iodine concentration. In Table 2, the mean bias ranges for low iodine concentrations were -0.34 mg/ml to -0.05 mg/ml and -0.16 mg/ml to - 0.06 mg/ml for large and small phantoms, respectively. In Table 3, the mean bias ranges for large and small phantoms were -0.44 mg/ml - 0.21 mg/ml and -0.35 mg/ml to -0.04 mg/ml, respectively. The difference in the mean bias measurements between individual acquisition combinations was minimal and all bias measurements were within the LOA. In Table 2, the widths of the LOA for low iodine concentrations were 0.32 mg/ml - 0.51 mg/ml and 0.12 mg/ml - 0.26 mg/ml for the large and small phantoms, respectively.

**Table 3.** Bland-Altman results calculated for high iodine concentrations (5, 10, 15 mg/ml), phantom sizes (small / large), and tube combinations (SS120, SS140, DS120, and DS140) over ultra-low radiation dose levels (CTDI_vol_ = 0.6, 0.8, 1.2 mGy).

| Phantom size | Acquisition mode | Bias [mg/ml] |  |  | Mean bias [mg/ml] | Limits of agreement |  |
| --- | --- | --- | --- | --- | --- | --- | --- |
|  |  | Iodine 5 mg/ml | Iodine 10 mg/ml | Iodine 15 mg/ml |  | Lower [mg/ml] | Upper [mg/ml] |
| Large | SS120 | 0.09 | 0.25 | -0.54 | -0.07 | -0.74 | 0.6 |
|  | SS140 | 0.09 | 0.51 | 0.03 | 0.21 | -0.21 | 0.62 |
|  | DS120 | -0.47 | 0.11 | -0.97 | -0.44 | -1.31 | 0.42 |
|  | DS140 | -0.17 | 0.51 | -0.12 | 0.07 | -0.54 | 0.69 |
| Small | SS120 | -0.27 | 0 | -0.77 | -0.35 | -0.97 | 0.27 |
|  | SS140 | -0.22 | 0.12 | -0.53 | -0.21 | -0.73 | 0.31 |
|  | DS120 | -0.2 | 0.06 | -0.63 | -0.26 | -0.82 | 0.3 |
|  | DS140 | -0.1 | 0.28 | -0.3 | -0.04 | -0.51 | 0.43 |

For medium radiation dose levels, iodine mean bias measurements ranged from -0.22 mg/ml to -0.12 mg/ml and -0.18 mg/ml to -0.12 mg/ml for large and small phantoms, respectively, considering low iodine concentrations (Table 4). In Table 5, the mean bias measurements ranged from -0.2 mg/ml - 0.31 mg/ml and -0.39 mg/ml to -0.26 mg/ml for the large and small phantoms, respectively. Only minor differences were observed between individual acquisition combinations, phantom configurations, and iodine concentration groups. Again, all bias measurements were within the designated LOA. The widths of the LOA for low iodine concentrations (Table 4) were marked from 0.19 mg/ml - 0.37 mg/ml and 0.04 mg/ml - 0.06 mg/ml for the large and small phantom, respectively. The corresponding widths of LOA for high iodine concentrations (Table 5) were noted from 1.0 mg/ml - 1.12 mg/ml and 0.69 mg/ml - 0.72 mg/ml.

**Table 4.** Bland-Altman results calculated for low iodine concentrations (0.2, 0.5, 1, 2 mg/ml), phantom sizes (small / large), and tube combinations (SS120, SS140, DS120, and DS140) over medium radiation dose levels (CTDI_vol_ = 6, 10, 15 mGy).

| Phantom size | Acquisition mode | Bias [mg/ml] |  |  |  | Mean bias [mg/ml] | Limits of agreement |  |
| --- | --- | --- | --- | --- | --- | --- | --- | --- |
|  |  | Iodine 0.2 mg/ml | Iodine 0.5 mg/ml | Iodine 1 mg/ml | Iodine 2 mg/ml |  | Lower [mg/ml] | Upper [mg/ml] |
| Large | SS120 | -0.07 | -0.2 | -0.28 | -0.29 | -0.21 | -0.38 | -0.04 |
|  | SS140 | 0.01 | -0.17 | -0.24 | -0.06 | -0.12 | -0.3 | 0.07 |
|  | DS120 | -0.07 | -0.24 | -0.31 | -0.26 | -0.22 | -0.4 | -0.05 |
|  | DS140 | -0.13 | -0.21 | -0.23 | -0.12 | -0.17 | -0.27 | -0.08 |
| Small | SS120 | -0.12 | -0.15 | -0.16 | -0.16 | -0.15 | -0.18 | -0.12 |
|  | SS140 | -0.13 | -0.11 | -0.11 | -0.13 | -0.12 | -0.14 | -0.1 |
|  | DS120 | -0.17 | -0.18 | -0.19 | -0.17 | -0.18 | -0.2 | -0.16 |
|  | DS140 | -0.17 | -0.17 | -0.15 | -0.13 | -0.16 | -0.18 | -0.13 |

**Table 5.**
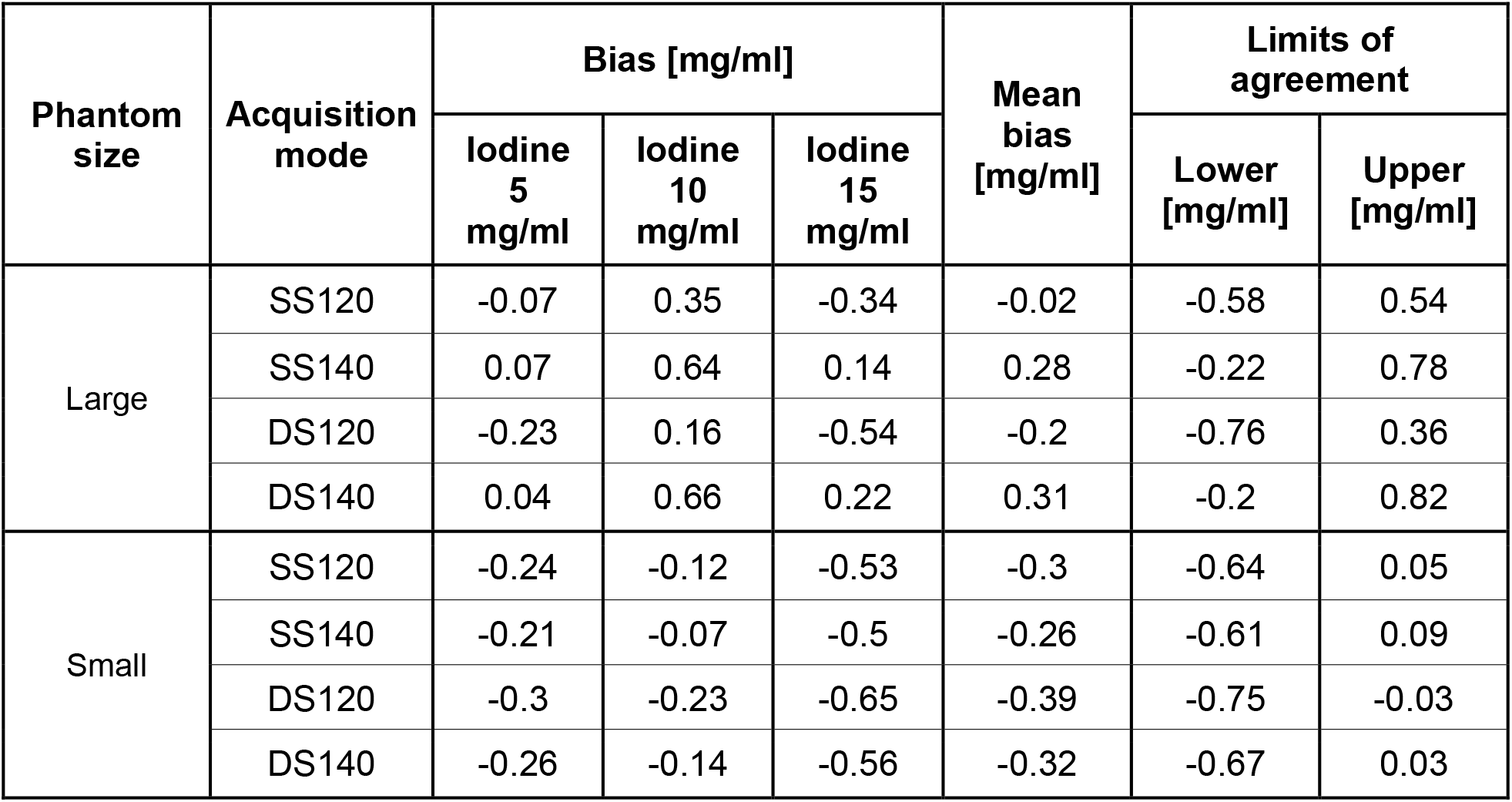
Bland-Altman results calculated for high iodine concentrations (5, 10, 15 mg/ml), phantom sizes (small / large), and tube combinations (SS120, SS140, DS120, and DS140) over medium radiation dose levels (CTDI_vol_ = 6, 10, 15 mGy).

| Phantom size | Acquisition mode | Bias [mg/ml] |  |  | Mean bias [mg/ml] | Limits of agreement |  |
| --- | --- | --- | --- | --- | --- | --- | --- |
|  |  | Iodine 5 mg/ml | Iodine 10 mg/ml | Iodine 15 mg/ml |  | Lower [mg/ml] | Upper [mg/ml] |
| Large | SS120 | -0.07 | 0.35 | -0.34 | -0.02 | -0.58 | 0.54 |
|  | SS140 | 0.07 | 0.64 | 0.14 | 0.28 | -0.22 | 0.78 |
|  | DS120 | -0.23 | 0.16 | -0.54 | -0.2 | -0.76 | 0.36 |
|  | DS140 | 0.04 | 0.66 | 0.22 | 0.31 | -0.2 | 0.82 |
| Small | SS120 | -0.24 | -0.12 | -0.53 | -0.3 | -0.64 | 0.05 |
|  | SS140 | -0.21 | -0.07 | -0.5 | -0.26 | -0.61 | 0.09 |
|  | DS120 | -0.3 | -0.23 | -0.65 | -0.39 | -0.75 | -0.03 |
|  | DS140 | -0.26 | -0.14 | -0.56 | -0.32 | -0.67 | 0.03 |

Figure 5 presents the effect of QIR across all radiation dose levels (CTDI_vol_ = 0.6 - 15 mGy) acquired using SS120 acquisition mode. RMSE values calculated over all (0.2 - 15 mg/ml), low (0.2, 0.5, 1, 2 mg/ml), and high (5, 10, 15 mg/ml) iodine concentrations are shown in three plots. Comparable RMSE values were calculated between individual iterative reconstruction levels across all radiation dose levels in all three cases. On average, overall RMSE values and those for low iodine concentrations were less than 0.3 mg/ml and 0.2 mg/ml, respectively. Although higher RMSE values were calculated for high iodine concentrations, the RMSEs were less than 0.4 mg/ml.

**Figure 5.**
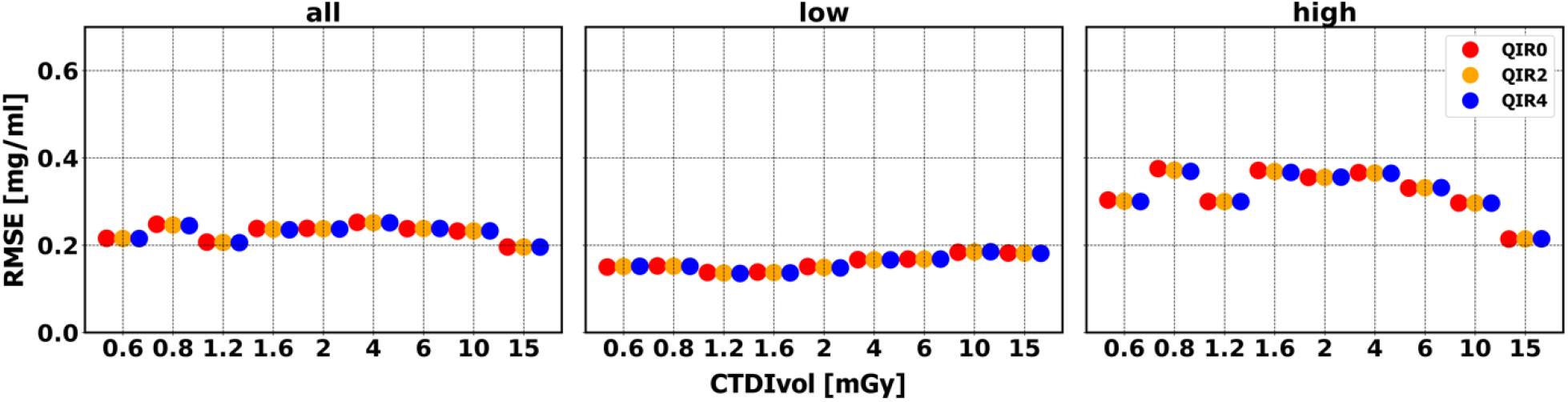
Comparison of RMSE [mg/ml] from different QIR levels (QIR0, QIR2, and QIR4) calculated over (**left**) all (0.2 - 15 mg/ml), (**middle**) low (0.2, 0.5, 1, 2 mg/ml), and (**right**) high iodine concentrations (5, 10, 15 mg/ml), across all radiation dose levels (CTDI_vol_ = 0.6 - 15 mGy) and obtained using SS120 acquisition mode. In each CTDI_vol_ column, QIR levels were color-coded and arranged by increasing QIR levels (QIR0, QIR2, and QIR4).

Figure 6 represents the influence of phantom configuration across all radiation dose levels (CTDI_vol_ = 0.6 - 15 mGy) acquired using SS120 acquisition mode. Computed RMSE values using all (0.2 - 15 mg/ml) and low (0.2, 0.5, 1, 2 mg/ml) were below 0.4 mg/ml while RMSE for high (5, 10, 15 mg/ml) iodine concentrations were less than 0.6 mg/ml. Minimal RMSE variations were evident from CTDI_vol_ = 1.2 - 6 and 15 mGy over all iodine concentrations. RMSE values calculated over high concentrations showed slight variations across all radiation dose levels except at CTDI_vol_ = 0.8 mGy. Finally, RMSE values for the small phantom improved with increasing CTDI_vol_ > 4 mGy.

**Figure 6.**
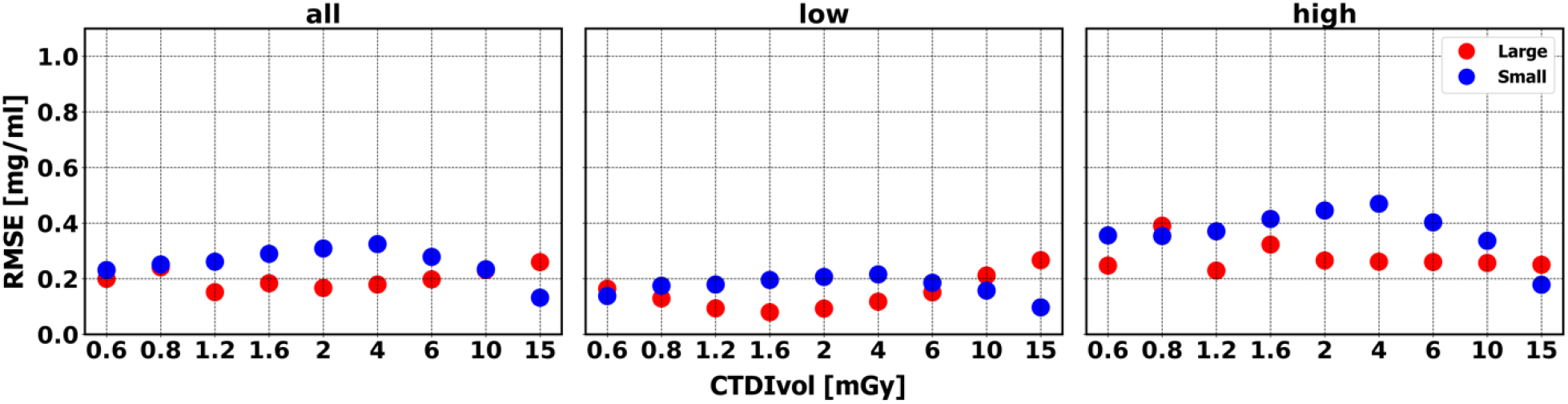
Comparison of RMSE [mg/ml] from different phantom sizes (small / large) calculated over (**left**) all (0.2 - 15 mg/ml), (**middle**) low (0.2, 0.5, 1, 2 mg/ml), and (**right**) high iodine concentrations (5, 10, 15 mg/ml), across all radiation dose levels (CTDI_vol_ = 0.6 - 15 mGy) and obtained using SS120 acquisition mode. In each CTDI_vol_ column, phantom sizes were color-coded.

## Discussion

In this study, we present an initial systematic evaluation of a recently introduced clinical DS-PCCT system for measurement of iodine concentrations. We investigated the influence of patient habitus, acquisition parameters, and iterative reconstruction modes on the accuracy of DS-PCCT iodine-specific images or iodine maps. Overall, our phantom study demonstrated accurate iodine quantification with minimal effect of patient habitus, tube voltage combinations, or iterative reconstruction on the iodine measurement accuracy of the DS-PCCT system, including at considerably lower than dose levels than used in the current clinical day-to-day routine. It was noted that despite the minimal increase in iodine bias, PCCT technology outperforms other spectral platforms due to its excellent ultra-low dose performance.

DS-PCCT measurements of various iodine concentrations demonstrated comparable accuracy compared to ground truth over a wide range of values (0.2 to 15 mg/ml). This included high measurement accuracy at low iodine concentrations (0.2 - 2 mg/ml) resulting in RMSE of less than 0.3 mg/ml across different phantom sizes, tube settings, and radiation dose groups. Based on data collected from an early prototype PCCT system, *Riederer et al*., reported similar iodine quantification performance^16^. Reliable quantification of iodine uptake, particularly of low iodine concentrations, may enable improved diagnosis including early tumor detection, differentiation, and staging. On the other hand, high iodine concentrations were associated with a slight increase in RMSE. *Lang et al*., on a research PCCT system, demonstrated similar observations^17^. All biases measured in this study were well within the LOA, though even lower measurement variability was seen with low iodine concentrations, the small phantom, and medium radiation dose levels.

Previous DECT studies reported a significant effect of patient habitus on the accuracy of iodine quantification at ultra-low radiation exposures^18,19^. The advent of PCD technology has improved the quality of diagnostic imaging of large patients significantly compared to EIDs^20,21^. In our quantitative evaluation, calculated RMSEs over all iodine concentrations for both phantom configurations remained consistently below 0.4 mg/ml, even for very low dose levels. Furthermore, only minimal RMSE discrepancies were seen between phantom configurations. These suggest accurate and consistent iodine concentration measurements regardless of the patient size.

Similar iodine quantification performance was observed across the iterative reconstruction levels at all radiation dose levels. These results indicate that the noise reduction, which is associated with QIR strengths, does not significantly affect iodine measurements. Similar observations have been made in phantom studies on available DECT systems^18,19,22^ and clinical studies with the DS-PCCT^12,23^.

We used a phantom model instead of patient data for analysis, which has several limitations. In clinical scans, iodine quantification can be affected by factors such as respiratory motion, metal artifacts, or morbid obesity. To overcome some of these limitations, multiple tube settings and phantom sizes were used in this study, but patients studies are still needed to account for these factors.

## Conclusion

Quantification of iodine uptake can assist in clinical diagnosis, such as distinguishing between a hyperdense cyst and a solid tumor. Accurate and reliable iodine density maps may eliminate the need for the unenhanced phase, which has served as a comparison for determining iodine uptake. It is anticipated that, in the future, accurate and precise iodine quantification will provide opportunities for the development of spectral-based biomarkers, such as those that may be employed to evaluate cancer treatments.

## Data Availability

All data produced in the present study are available upon reasonable request to the authors

## Abbreviations

CM: contrast media
CT: computed tomography
HU: Hounsfield units
PCCT: photon-counting computed tomography
PCD: photon-counting detector
EID: energy-integrating detectors
DECT: dual-energy computed tomography
DS: dual source
SS: single source
CTDI_vol_: CT dose index
QIR: quantum iterative reconstruction
VMI: virtual monoenergetic image
ROI: region of interest
RMSE: root mean square error
BA: Bland-Altman
LOA: limits of agreement

## Acknowledgment

We acknowledge support through the National Institutes of Health (R01EB030494) and Siemens Healthineers.

